# Feasibility study of a novel co-designed online training program for circus coaches working with preschool-aged children born preterm

**DOI:** 10.1101/2023.05.24.23290505

**Authors:** Free Coulston, Rachel Toovey, Kath Sellick, Rheanna M Mainzer, Loni Binstock, Alicia Spittle

**Author notes:** Corresponding author: Free Coulston - Postal address: Department of Physiotherapy, The University of Melbourne, Level 7 Alan Gilbert Building, 161 Barry St, Carlton, Victoria 3053, Australia.

## Abstract

**Purpose:** Providing specialised training to community-based physical activity instructors (such as circus coaches) has been identified as a potential strategy to increase participation for preschool-aged children born preterm. The objective of this study was to determine the feasibility of a novel co-designed training program “CirqAll: professional development for circus coaches” (CirqAll:PD), which aimed to increase coaches’ knowledge, skills, and confidence in working with children born preterm.

**Materials and methods:** CirqAll:PD consisted of 10-hours of online self-directed content and four 90-minute online workshops completed over four weeks. Recruitment capability, acceptability, implementation fidelity and limited efficacy testing were evaluated using a case series design.

**Results:** Fifty-one circus coaches were enrolled, and 27 completed CirqAll:PD. Reasons for attrition were primarily related to the Theoretical Framework of Acceptability’s (TFA) concept of burden. All 27 coaches indicated that CirqAll:PD was acceptable (TFA questionnaire). Overall intervention fidelity was high (high adherence to planned delivery, moderate adherence to dosage, and excellent participant responsiveness). Limited efficacy testing revealed positive trends regarding coaches’ knowledge, skills, and confidence (Determinants of Implementation Behaviour Questionnaire).

**Conclusions:** These results support the feasibility of CirqAll:PD. Adaptations to reduce attrition and burden on participants are required prior to further testing.

## Introduction

Ensuring healthy lives and promoting well-being for all ages is one of the United Nations Sustainable Development Goals [1]. Adequate levels of physical activity are crucial for good health [2] and inadequate physical activity is a major risk factor for chronic disease [3]. During childhood, physical activity is essential for growth and development [4]. Participation in physical activity positively impacts children’s health by strengthening the skeletal, cardiovascular, and immune systems and improving coordination, balance, posture, and flexibility. Furthermore, physical activity is also an important part of play and learning, and helps children develop language, social, and communication skills with peers [5–7].

Participation in physical activity in the preschool years (ages 3-5 years) is particularly important given the development of fundamental motor and social skills occurring at this age, as well as the development of lifelong physical activity patterns [8,9]. However, only six in ten preschool-aged children meet the Australian physical activity guidelines [10]. Preschool-aged children who were born preterm (less than 30 weeks’ gestation), participate in even less physical activity than children who were born at term (>37 weeks’ gestation) [11]. Preterm birth increases the likelihood of motor impairments, including cerebral palsy (CP) and developmental coordination disorder (DCD), which have been shown to affect participation in physical activity [11–15].

Participation in age-appropriate life activities such as community-based physical activity programs, is a key concept of child development [9]. However, preschool-aged children born preterm have been shown to have reduced community participation compared to term-born children [16]. Many opportunities for physical activity at the preschool age occur in community-based settings, such as kinder gym, preschool circus, or dance classes. As this is a formative age for both fundamental physical and social skill development, prioritising interventions that may increase physical activity participation for preschool-aged children born preterm is essential [2,17–19].

A recent mixed-methods study conducted by the authors explored the needs and preferences of key stakeholder groups regarding the participation of preschool-aged children born preterm in community-based physical activities [20]. The stakeholder groups included parents of children born extremely preterm (less than 28 weeks’ gestation), paediatric clinicians, and circus coaches. Circus was chosen as the community-based physical activity of interest due to its potential to improve physical and social outcomes for varied paediatric populations, including those with CP and developmental delay [21]. Stakeholders proposed that a key strategy to increase participation for children born preterm is for providers (such as coaches and organisations) to be understanding of the preterm experience and have specialised knowledge in inclusion and optimising developmental outcomes. Furthermore, parents’ heightened vigilance, risk aversion and the diverse needs of their children requires coaches to tailor programs to support initial engagement and ongoing participation. A further study by the authors utilised a co-design process to develop a circus-based intervention that included professional development training for coaches in these areas [22]. This novel co-designed training program is titled *“CirqAll: professional development for circus coaches*” (CirqAll:PD).

Where a novel intervention such as CirqAll:PD is proposed, it is important to assess its feasibility (can it work?) before proceeding to a larger efficacy trial (does it work?) [23,24]. Feasibility testing can determine whether an intervention should be recommended for efficacy testing and *“identify not only what—if anything—in the research methods or protocols needs modification but also how changes might occur”* [23, p. 2]. This can prevent wasted resources such as funding, participant and researcher time, and assist in prioritizing interventions with the greatest likelihood of being efficacious [23,24]. This case series feasibility study aims to answer the research question *“Is CirqAll:PD a feasible intervention?”* by evaluating:

- recruitment capability,
- acceptability of the intervention to circus coaches,
- implementation fidelity, and
- limited efficacy testing on a key outcome of interest: coaches’ knowledge, skills and confidence in working with children born preterm.

## Materials and methods

### Research design

A case series design [25] was chosen for this study. Case series are recommended for feasibility trials [23] and have been utilised in another study looking at a novel dance-based physical activity intervention [26]. This study received ethical approval from the Royal Children’s Hospital Human Research and Ethics Committee (Ethics approval number: HREC/84753/RCHM-2022) and is registered with the Australian New Zealand Clinical Trials Registry on 5th April 2022 (trial registration number: ACTRN12622000532707p).

### Participants

This study aimed to recruit a sample size of 42 coaches to obtain a 95% confidence interval of +/- 10% around an acceptability percentage estimate of 90% (i.e., expected 95% confidence interval 80% to 100%). Circus coaches were recruited via convenience sampling by contacting Australian circus schools and providing them with study flyers to distribute to their coaches. Study flyers were also distributed via social media [27], and via an email update to interested and consenting coaches who had participated in a prior study by the authors [20]. Coaches needed to meet the following eligibility criteria to participate in the study:

- be 16 years or older at the time of study enrolment,
- have taught more than 30 hours of circus skills to children aged 3-5 years old,
- have taught circus skills to children aged 3-5 years old in the past five years in Australia,
- have access to the internet and a device that connects to the internet,
- be able to comprehend both written and spoken English.

Coaches accessed the plain language statement and expressed interest in participating via an online REDCap survey displayed on the study flyer. REDCap (Research Electronic Data Capture hosted at Murdoch Children’s Research Institute, Melbourne, Australia) is a secure web platform for managing online databases and surveys [28,29]. Coaches were then contacted by a research assistant unknown to them (L.B.), and screening and informed consent discussions were undertaken. Eligible coaches were emailed an e-consent form to electronically sign via REDCap. Coaches under 18 years of age had their parent present during the informed consent discussion, and both provided e-consent. Coaches who completed the intervention and outcome measures were provided with a $200AUD e-gift voucher to acknowledge the time commitment required.

### Intervention description

CirqAll:PD was co-designed by parents of children born preterm, clinicians, circus coaches and researchers as described in [22], and co-produced by members of the co-design team and other experts (Figure 1). CirqAll:PD trains coaches on how to deliver inclusive classes to enhance participation of children born preterm, and its aim is to increase circus coaches’ knowledge, skills, and confidence in working with children born preterm. An online delivery method was chosen as the intervention was developed during COVID-19 restrictions which prohibited face-to-face teaching. The literature supports an online approach, citing increased accessibility and flexibility with comparable learning outcomes [30,31].

**Figure 1:**
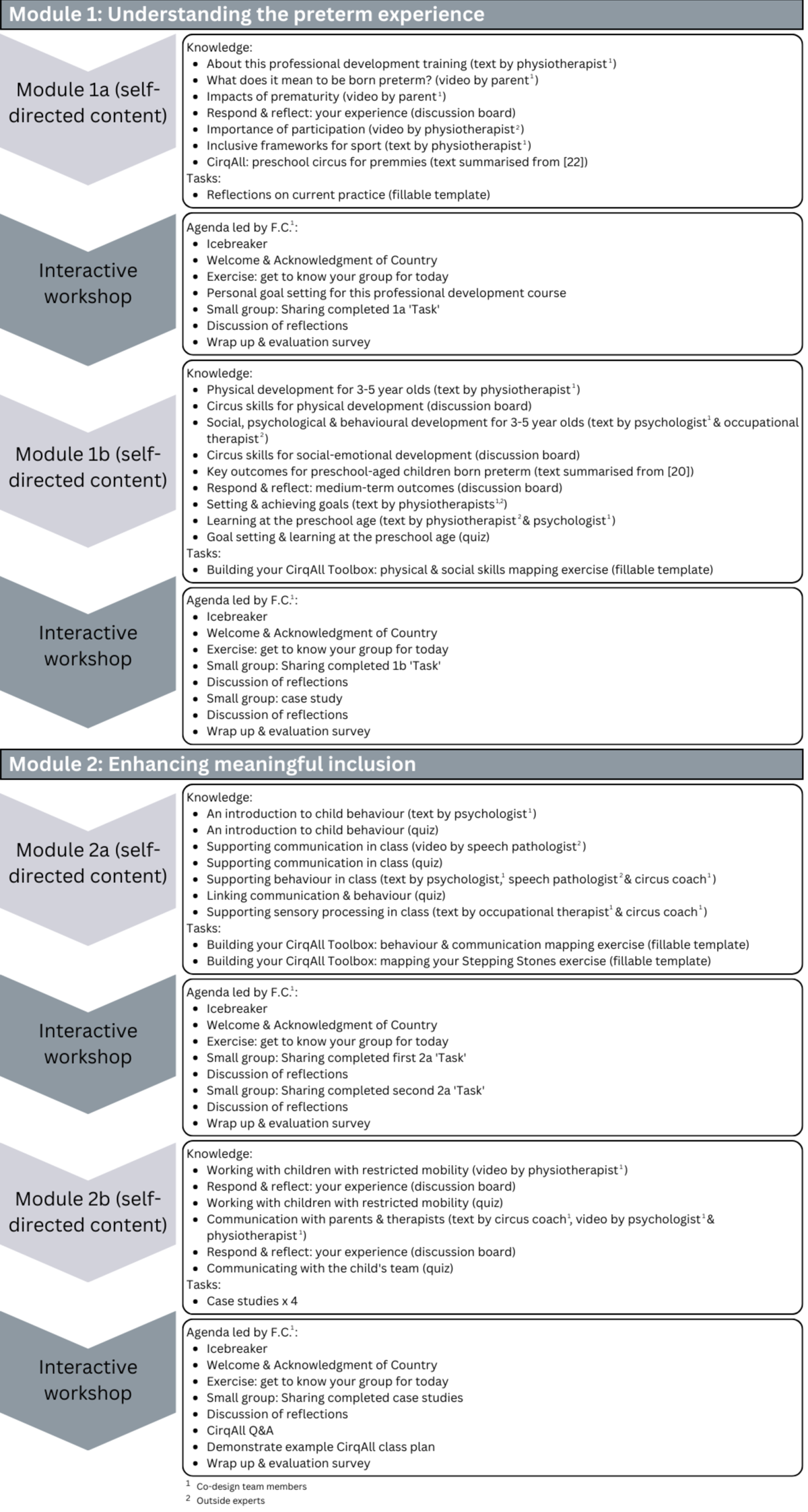
Structure and content of CirqAll:PD. Figure 1 Alt Text: A map showing two modules of the training, with four components and their content listed for each module.

A flipped classroom approach was applied [32] whereby coaches completed a series of online self-directed modules (via FutureLearn, hosted by The University of Melbourne), with each module followed by an interactive workshop (via Zoom) to apply and extend the concepts introduced. Interactive workshops were delivered twice each, and recorded, to offer increased flexibility for participants. Modules took approximately 2.5 hours to complete, and workshops ran for 90 minutes, resulting in a total intervention time of 16 hours over four weeks.

### Data collection

The primary objective of this study was to determine the feasibility of CirqAll:PD *(can it work?)* through evaluating recruitment capability, intervention acceptability, intervention fidelity and limited efficacy testing [23,24]. The study procedure and outcome assessment timeline are illustrated in Figure 2.

**Figure 2:**
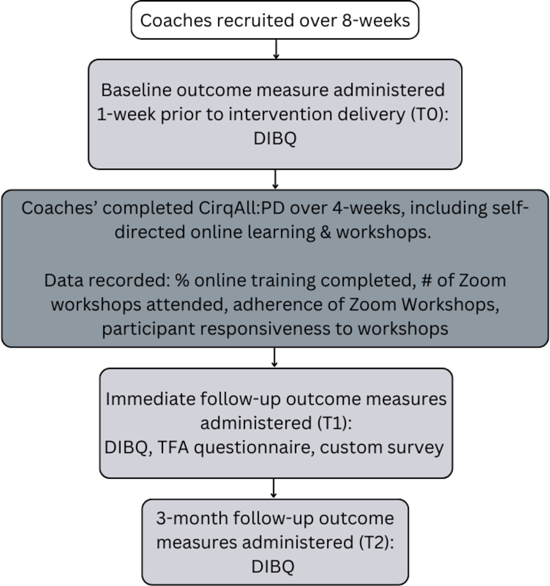
Study timeline. Figure 2 Alt Text: A flowchart showing key elements of the study, including timing of recruitment, delivery of intervention and application of outcome measures.

### Evaluation of recruitment capability

Recruitment capability is defined as the ability to recruit sufficient appropriate participants in an acceptable timeframe [24]. To assess the capability of the recruitment strategy described above, the numbers of coaches expressing interest were compared with the numbers of enrolled coaches. The duration of the recruitment period (the time from enrolment opening to the final participant enrolled) and any challenges to recruitment were recorded to provide further insight into the suitability of the recruitment strategy. Recruitment success was defined as enrolling 42 eligible circus coaches within eight weeks.

### Evaluation of intervention acceptability

Acceptability is *“a multi-faceted construct”* that describes the appropriateness of the intervention from the perspective of the participants [33, p. 1]. Acceptability of CirqAll:PD was assessed via the Theoretical Framework of Acceptability (TFA) questionnaire [34]. This was distributed to coaches via REDCap after completion of the intervention. Acceptability was also assessed through retention and attrition data, which was recorded at each outcome measure time point described in Figure 2. Considering these factors, overall acceptability of CirqAll:PD was defined by the authors as:

a. 90% of coaches scoring item 8 of the TFA questionnaire (overall acceptability question) as 4 or higher, and
b. <20% of coaches withdrawing citing reasons related to acceptability constructs (as per definitions for each construct from the TFA).

### Evaluation of intervention fidelity

Fidelity assessment of intervention implementation should measure process (e.g., participant responsiveness) and structure (including both dosage and adherence) [35]. Adherence to intervention protocol was quantified using a checklist of agenda items for the Zoom workshops. Agenda items were marked by the workshop facilitator (F.C.) as ‘Present’ (score of 2), ‘Attempted’ (score of 1) or ‘Absent’ (score of 0). The overall adherence score was calculated as a percentage of the total possible score. Intervention dosage was quantified by recording the number of coaches who completed ≥90% of the online self-directed content, and the number of interactive workshops attended. Participant responsiveness was measured using a 5-point Likert scale, where coaches anonymously rated their involvement during each workshop on an online REDCap survey, from 1 = not at all involved/engaged to 5 = very involved/engaged. In the absence of clear definitions in the literature [36], high fidelity of CirqAll:PD was defined by the authors as:

a. 75% of coaches completing ≥90% of the online learning, and
b. 75% of coaches attending ≥3 Zoom workshops (75%), and
c. The fidelity checklist for Zoom workshop agenda items scoring 75% or greater for each workshop, and
d. Coaches scoring ≥3 on the involvement scale, in ≥3 workshops.

### Evaluation of limited efficacy testing

Insight into the efficacy of CirqAll:PD was assessed using the Determinants of Implementation Behaviour Questionnaire (DIBQ) [37]. The DIBQ is a validated questionnaire that can provide information on whether CirqAll:PD is effective in increasing coaches’ knowledge, skills and confidence in working with children born preterm (the goal of the program) [38]. The DIBQ was administered electronically via REDCap at baseline, immediately after completing the four-week intervention period, and three-months later to assess changes in outcome measures.

### Data Analysis

Analysis using descriptive statistics was carried out in STATA, version 14 (Stata Corp, College Station, TX, USA) by F.C. and R.M. in consultation with the broader research team.

## Results

### Evaluation of recruitment capability

The recruitment strategy was successful as 51 coaches were consented and enrolled within eight-weeks (Figure 3). This represented 83.6% of all coaches who expressed interest (51/61) and exceeded the proposed sample size of 42 (see Methods). Reasons for non-enrolment are recorded in Figure 3. The sample characteristics of enrolled coaches are described in Table 1.

**Figure 3:**
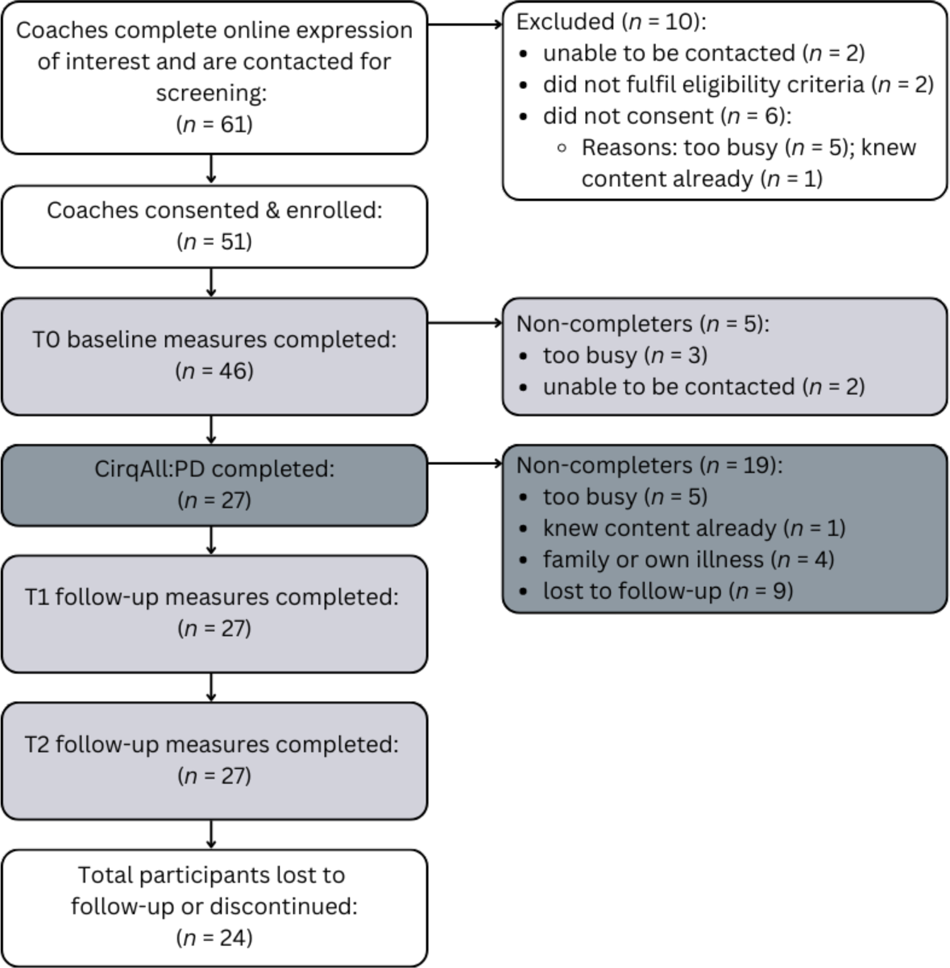
Flow of participants through the study. Figure 3 Alt Text: A flowchart showing the number of participants retained at each stage of the study and the reasons why participants dropped out or withdrew.

**Table 1:**
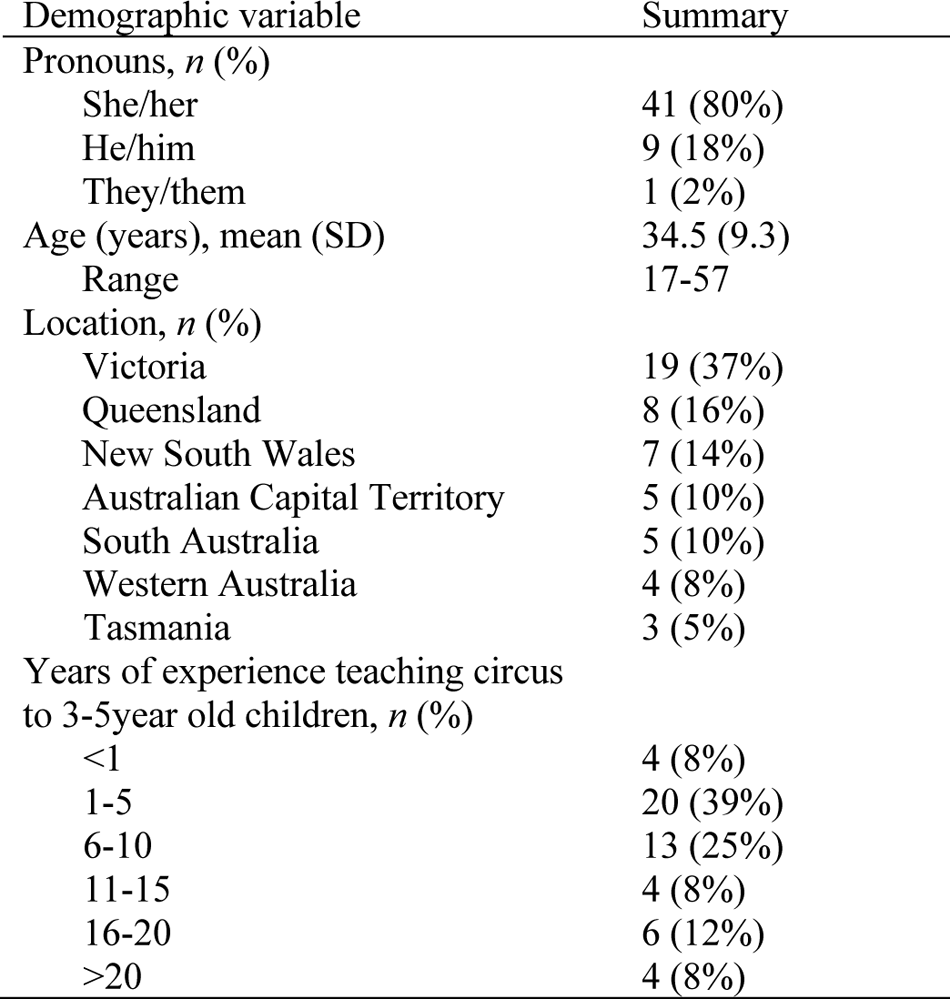
Demographics of *n* = 51 enrolled coaches.

| Demographic variable | Summary |
| --- | --- |
| Pronouns, $n$ (%) | |
| She/her | 41 (80%) |
| He/him | 9 (18%) |
| They/them | 1 (2%) |
| Age (years), mean (SD) | 34.5 (9.3) |
| Range | 17-57 |
| Location, $n$ (%) | |
| Victoria | 19 (37%) |
| Queensland | 8 (16%) |
| New South Wales | 7 (14%) |
| Australian Capital Territory | 5 (10%) |
| South Australia | 5 (10%) |
| Western Australia | 4 (8%) |
| Tasmania | 3 (5%) |
| Years of experience teaching circus to 3-5year old children, $n$ (%) | |
| <1 | 4 (8%) |
| 1-5 | 20 (39%) |
| 6-10 | 13 (25%) |
| 11-15 | 4 (8%) |
| 16-20 | 6 (12%) |
| >20 | 4 (8%) |

### Evaluation of intervention acceptability

Acceptability data was varied due to the combination of significant attrition, but high acceptability scores reported by those who were retained. Prior to the intervention beginning, five enrolled coaches failed to complete the baseline measures (reasons detailed in Figure 3) and were excluded from the study. Forty-six coaches completed the baseline measures, but only 27 coaches (58.7% of remaining participants) completed the intervention. For coaches who withdrew due to being too busy or due to illness (*n* = 9), this might align with the TFA construct of burden, which describes *“the perceived amount of effort that is required to participate in the intervention (e.g., participation requires too much time or expense, or too much cognitive effort, indicating the burden is too great)”* [33, p. 8]. For the coach who indicated the content of the intervention was known to them already, their reason for withdrawing may align with the TFA construct of perceived effectiveness (e.g., this intervention which aims to increase coaches’ knowledge and confidence is unlikely to achieve its purpose in this case). Therefore ≥20% (21.7%; 10/46) of coaches withdrew citing reasons related to acceptability constructs. Coaches who completed the intervention and those that did not were comparable regarding collected demographic characteristics (Appendix A).

Of the 27 coaches who completed the intervention, all (100%) scored item 8 of the TFA (General Acceptability Score [34]) as either acceptable or completely acceptable (Figure 4). [Figure 4 near here] Graphs illustrating coaches’ responses to the remaining TFA items are presented in Appendix B. Most coaches (25/26) understood the CirqAll:PD intervention and its purpose, and perceived that CirqAll:PD was effective in its aim (24/27) (Item 5, Intervention Coherence; Item 4, Perceived Effectiveness).

**Figure 4:**
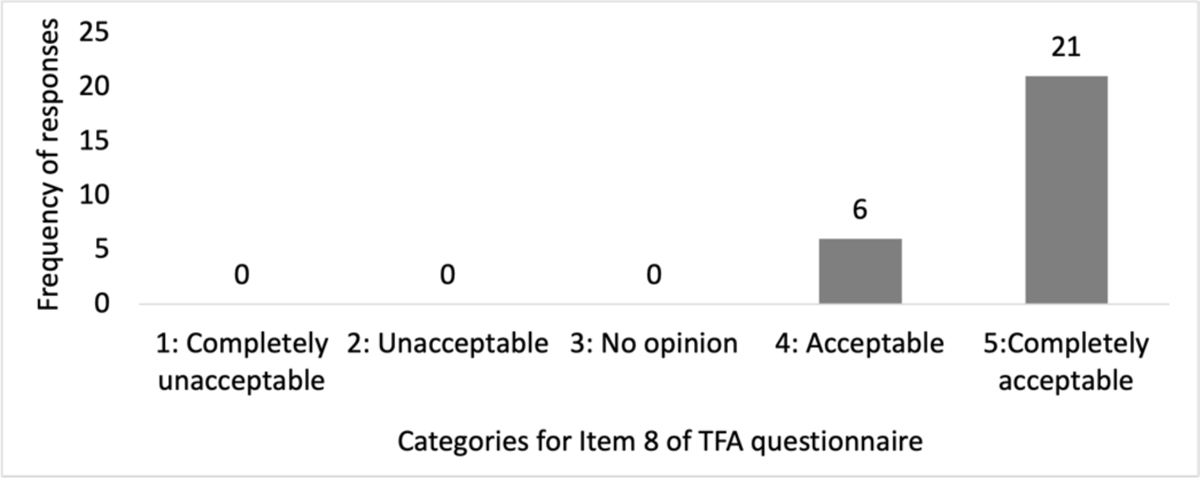
Participant responses to the question *“How acceptable was the Coach Training to you?”* [34]. Figure 4 Alt Text: Bar graph showing six participants indicating their answer as acceptable, and 21 participants as completely acceptable.

Six coaches reported that participating in CirqAll:PD required a lot of effort (6/27), and 9/27 coaches felt that participating in the intervention interfered with their other priorities (Item 2, Burden; Item 7, Opportunity Costs). However, 18/27 coaches felt there was a little effort involved and 11/27 coaches did not feel that CirqAll:PD interfered with their other priorities. Item 6, Self-efficacy, indicated that 25/27 coaches were confident they could engage in the intervention and felt that CirqAll:PD was a good fit with their personal values (Item 3, Ethicality).

Item 1, Affective Attitude, indicated that coaches felt positively towards CirqAll:PD (26/27). Furthermore, when asked in the follow-up custom survey what elements of CirqAll:PD they liked the most, coaches appreciated the discussions with peers (*n* = 13), gaining new knowledge (*n* = 9), well-organised accessible information (*n* = 9) and the varied formats in the CirqAll:PD delivery (*n* = 6) (Table 2).

**Table 2:** Participant responses to the question “Please let us know what you liked most about this professional development course”.

| Participants favourite CirqAll:PD elements | Number of responses ($n$) |
| --- | --- |
| Discussions with peers in collaborative workshops & on discussion boards | 13 |
| Gaining new knowledge, access to resources/ downloads | 9 |
| Well-organised, accessible information | 9 |
| Varied delivery/ formats (e.g., case studies, quizzes, workshops, videos, text) | 6 |
| Practical and relevant application | 4 |
| Reinforcing current practice | 2 |
| Online delivery | 1 |
| Workshop facilitator | 1 |

### Evaluation of intervention fidelity

#### Structure

Regarding dosage, 63% of coaches (17/27) completed ≥90% of the online self-directed content. Two additional coaches completed ≥50% of steps, and 29.6% (8/27) of coaches completed less than 50% of the self-directed content. Only three coaches attended all four interactive workshops synchronously, however 79% of coaches who missed one or more workshops (19/24) watched the recording/s asynchronously (Figure 5). Table 3 shows the reasons coaches did not attend workshops, or complete online learning items, as well as suggestions to improve the course.

**Figure 5:**
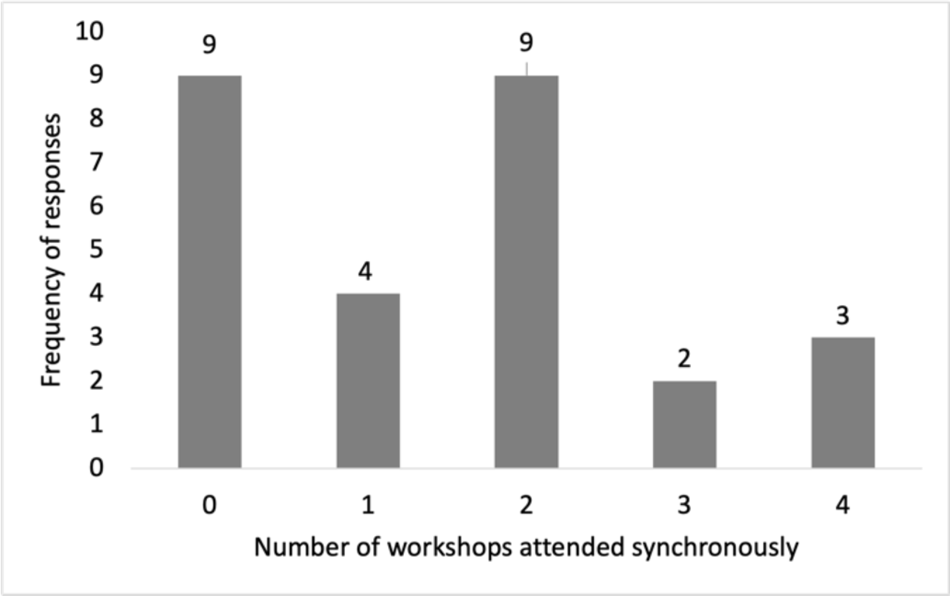
Participant responses to the question *“How many workshops did you attend?”.* Figure 5 Alt Text: Bar graph showing the numbers of synchronous workshops attended by participants.

**Table 3:** Reasons given by participants for non-attendance and suggestions for improvements.

| Reasons | Non-attendance at workshops (*n*) | Non-completion of any online self-directed learning (*n*) | Suggestions for improvements (*n*) |
| --- | --- | --- | --- |
| Schedule conflict/s (e.g., work or family commitments) | 20 |  | More options of times for workshops (4)<br>Don't need workshops (2) |
| Travelling/ Overseas | 4 |  |  |
| Health-related (self or family members) | 5 | 1 |  |
| Overcommitted/ too busy/ life stresses | 3 | 5 |  |
| Not enough time to complete |  | 6 | Access to recordings and materials for longer (1)<br>Course open for longer (4) |
| Website not user friendly |  | 3 | Print out of learning materials (eg. PDF booklet) (5) |
| Other |  |  | Eligibility criteria felt limiting (1)<br>Some survey questions difficult to follow (1) |

The overall adherence of the interactive workshops to the planned agenda was high (94.9%), with a score of 148 out of a possible 156. Four agenda items received a score of zero as the group numbers were smaller than anticipated and so a small group activity was completed all together instead.

#### Process

Thirty-nine anonymous survey responses were captured from coaches attending the interactive workshops (Figure 6), and all 39 responses indicated that coaches were either almost always, or mostly engaged, indicating excellent participant responsiveness.

**Figure 6:**
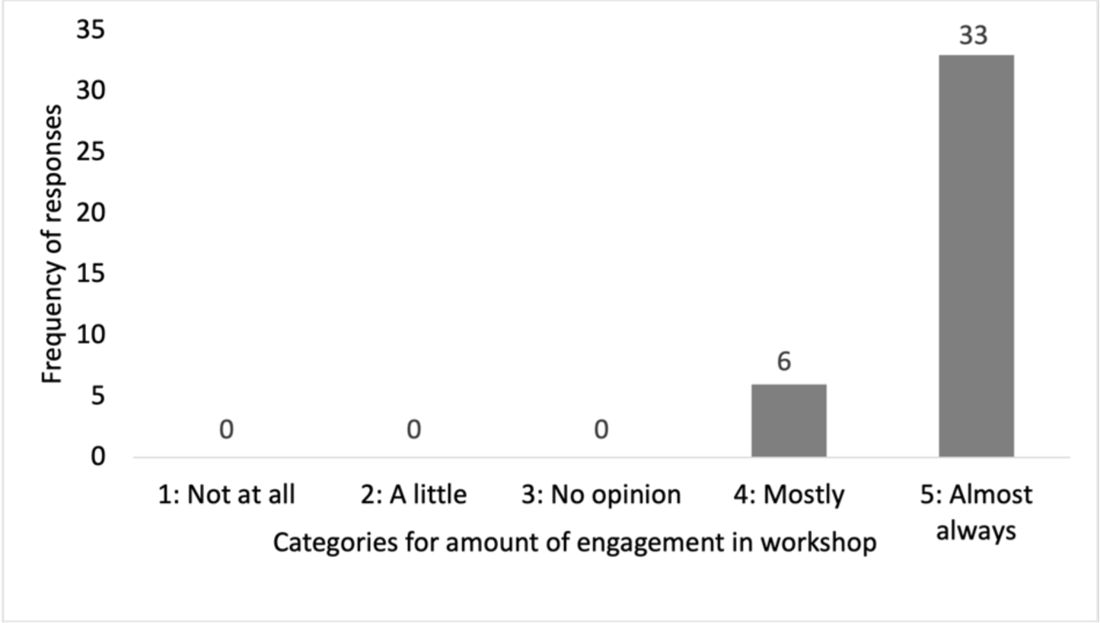
Participant responses in workshops to the statement *“Please rate your engagement in this workshop”.* Figure 6 Alt Text: Bar graph showing six responses from participants indicating mostly engaged in workshops, and 33 responses indicating almost always engaged.

### Evaluation of limited efficacy testing

Coaches who completed the intervention (*n* = 27) showed improvements on all domains of the DIBQ from baseline to post-intervention, with improvements retained at the three-month follow-up (Appendix C).

Changes in the specific determinants of knowledge, skills, and beliefs about capabilities (confidence) in working with children born preterm can be seen in Figure 7. Overall, there was a pattern to suggest that determinants of implementation related to these three domains of interest were increased.

**Figure 7:**
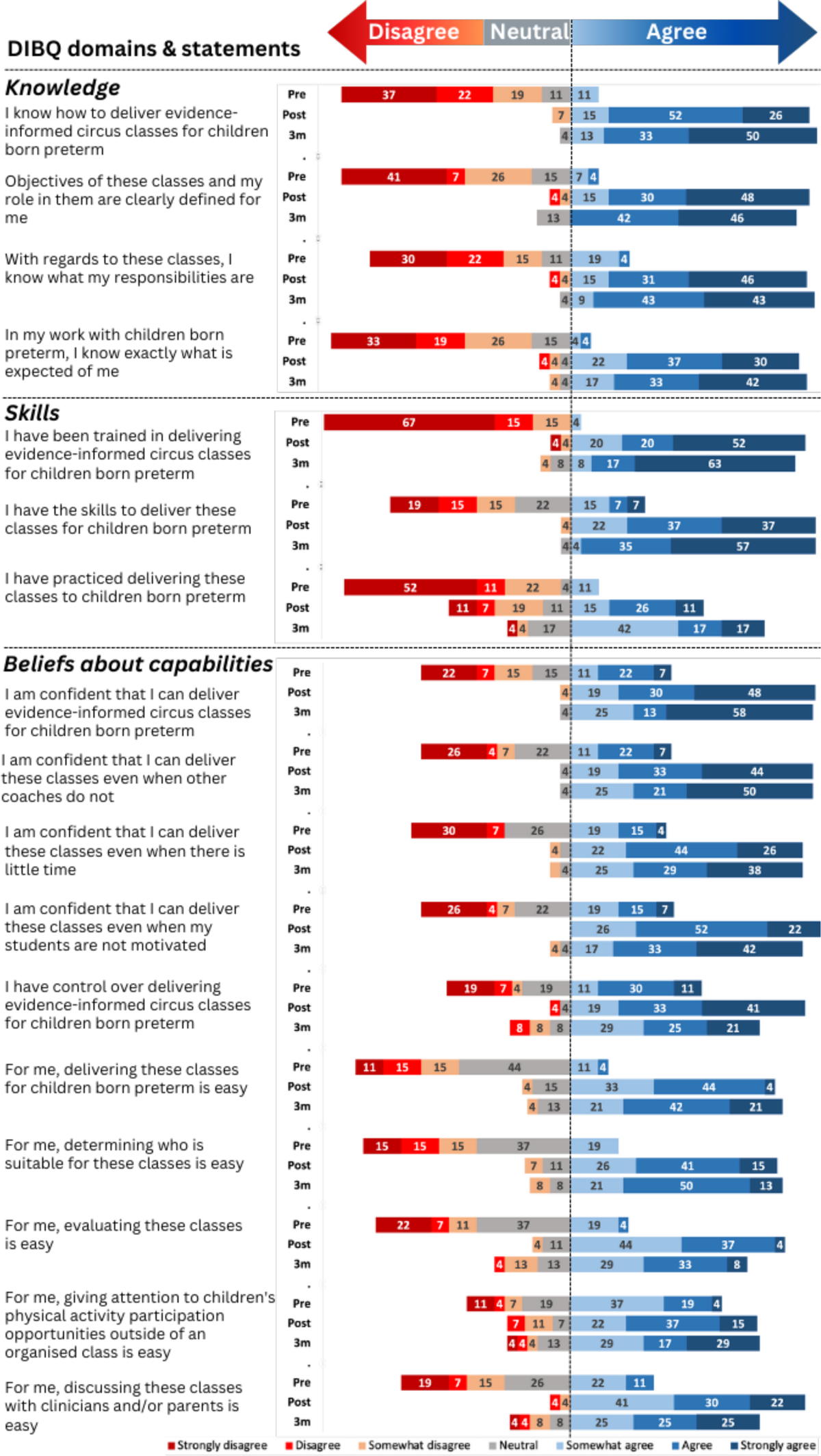
Results for the determinants of Knowledge, Skills and Beliefs about Capabilities [34] at baseline (pre), immediately following the intervention (post) and at the three month follow-up (3m). Figure 7 Alt Text: A colorful bar graph showing that coaches had improved from baseline to immediately after the training, at that those improvements had been retained at 3-months.

## Discussion

This study was guided by the research question *“Is CirqAll:PD a feasible intervention?”* Evaluation of recruitment capability, intervention fidelity, acceptability, and limited efficacy testing indicates that CirqAll:PD is feasible with some modifications required to reduce attrition.

Recruitment capability was adequate and intervention fidelity was high in the present study. These aspects of the study are feasible and do not require modification before proceeding to a larger scale trial. Likewise, limited efficacy testing revealed positive trends regarding improvements in, and retention of, coaches’ knowledge, skills, and confidence in working with children born preterm. Therefore, the content of CirqAll:PD is appropriate for the aim of the intervention, and should also be carried forward to a larger trial. This is supported by coaches who completed CirqAll:PD, liking the intervention (26/27) and rating it as acceptable (6/27) or completely acceptable (21/27) (Items 1 & 8, TFA questionnaire [34]).

However, as attrition of participants was also high (47% (23/51)) some modifications to reduce burden may be required to reduce attrition [33]. As attrition may be higher in online learning compared to face-to-face delivery (as discussed in literature reviews conducted by Stone [39] and Su & Waugh [40]) understanding how to improve retention from participants’ perspectives is key. Suggestions from CirqAll:PD participants (Table 3) indicate that extending the time to complete the course material, as well as providing the material in printed form, may improve retention. Suggestions regarding offering more times for the workshops to be conducted may not be feasible from a study perspective, as the workshops were already repeated in, and out, of business hours and also recorded. Two participants suggested that the workshops may not be an essential part of the intervention. However, feedback from participants regarding their favourite part of the intervention was primarily around the chance to openly collaborate with other like-minded coaches during the workshops (Table 2).

Although CirqAll:PD is the first online professional development course seeking to improve the knowledge, skills and confidence of circus coaches in working with children born preterm, there are a few other studies reporting online training for providers of interventions. Baruah et al. [41] undertook a randomised controlled trial looking at the feasibility of an online training program for carers of people with dementia. They reported a retention rate of 36%, with 45% of retained participants (*n* = 13) not completing a single online lesson [41]. In comparison, CirqAll:PD’s retention rate was 53%, and 70% of coaches completed >50% of the online training steps. Baruah et al. [41] did not collect reasons for attrition however, the authors surmise that a dislike of online delivery, no real need for the intervention, or the high proportion of males who signed up for the program (when higher rates of females are reported to be caregivers) may have played a part in the high attrition rates. These reasons are unlikely to apply to CirqAll:PD, as a prior study showed that online intervention delivery was acceptable to coaches and that coaches identified a need for this intervention [20]. Furthermore, the demographics of enrolled coaches (Table 1) correlate well with the intended recipients of the CirqAll:PD intervention, with no major differences in collected characteristics between completers and non-completers (Appendix A). It is possible that there were factors not explored that did affect completion. For example, as 80% of participants enrolled in this study identified with the pronouns she/her, and mean age was 34.5 years, caring responsibilities may have been such a factor. Although online part-time learning presents an opportunity to engage in education alongside other commitments and thus impacts educational access positively [39], *“women are likely to be carrying an additional caring load, which by necessity, is time and energy consuming and frequently needs to be prioritised over study”* [42, p. 105]. Caring responsibilities were also exacerbated during the COVID-19 pandemic, disproportionately affecting women and contributing to exhaustion and burnout [43].

An integrative review explored facilitators and barriers related to the feasibility of online training interventions for community volunteers supporting older adults [44]. Facilitators of feasibility included: relevant topics and high engagement to prevent attrition [44]. CirqAll:PD’s topics (Figure 1) are likely to be relevant given that they were derived through a stakeholder engagement process and refined in a co-design and co-production process with circus coaches. Hill et al. [44] describe high engagement as instructor enthusiasm, diversity in training formats, and having a participatory component. Participants in CirqAll:PD also described an appreciation of the varied formats, and peer collaboration through discussion boards and collaborative workshops (Table 2). However, as only one participant mentioned the workshop facilitator as being their favourite element, perhaps having multiple or varied facilitators could increase retention. The review by Hill et al. [44] also found that providing handbooks (e.g., hardcopy versions of the training) was a strategy used by several online interventions. This was also a strategy suggested by the CirqAll:PD participants, and therefore is recommended as a modification to CirqAll:PD prior to efficacy testing.

Barriers to online training feasibility identified in Hill et al.’s review included high workload of participants and time elapsed between the training and its application [44]. Five of the coaches who expressed interest in CirqAll:PD but declined to enrol cited being too busy, as did a further 33% (8/24) of non-completers. This appears to be a key factor in attrition in the current study, and although complex, strategies that account for busy professional and personal lives should be explored. Suggestions from CirqAll:PD participants including extending the time for intervention completion may assist in mitigating this factor. Another strategy (not proposed by participants) may be to reduce the amount of content in the course, and thus the time-burden required to complete it. Although this strategy may improve retention, reducing the depth or breadth of the content may result in less favourable improvements in coaches’ determinants of implementation behaviour (e.g., knowledge, skills & confidence).

Ongoing, sustainable follow-up support is proposed as a strategy to promote self-efficacy for implementing interventions following online training and may assist in coaches feeling more supported throughout the intervention, resulting in higher retention [20,44]. Interestingly, even without this follow-up support, the limited efficacy data collected during this feasibility study demonstrated that coaches’ determinants of implementation behaviour continued to increase over the three-month follow-up period.

In summary, the results of this study support the feasibility of CirqAll:PD. Prior to efficacy testing in a larger-scale trial, modifications to improve participant retention should be considered. These modifications may include extending the time to complete the course material, providing the material in printed form, varied workshop facilitators, and ongoing sustainable follow-up support.

### Strengths and limitations

A strength of this study is that CirqAll:PD was co-designed with circus coaches, parents of children born preterm, and paediatric clinicians. This involvement of key stakeholder groups improves the likelihood of intervention uptake and relevance to end-user needs [45,46]. Although limitations of this study include the convenience sampling approach and the attrition, which make it challenging to generalise results from the coaches who completed the program, the use of standardised tools with psychometric properties to capture efficacy data enhances its rigor. Furthermore, the evidence-based design of the feasibility study supports the progression of CirqAll:PD to proceed to larger-scale efficacy testing pending some modifications.

## Conclusion

Understanding if this novel co-designed training program *can* work is an important step prior to conducting a large-scale efficacy trial [23,24]. The results of this feasibility study suggest that CirqAll:PD be recommended for efficacy testing, after undergoing modifications to reduce attrition.

## Data Availability

The data that support the findings of this study are available from the corresponding author, FC, upon reasonable request.

## Acknowledgments

We would like to thank everyone who contributed their knowledge and expertise towards the content development of CirqAll:PD, including Co-Design Team members Loni Binstock, Caroline Keating, Isaac Fletcher, Kimberley Attard, and outside experts Kate Cameron, Nyssa Johnson and Ruby Rowat. Thanks also go to Alexander Kimp for assisting with the Learning Design in FutureLearn. This study will contribute to F.C.’s PhD candidature through The University of Melbourne.

## Appendix A: Comparison of collected demographic characteristics for coaches who completed the intervention, and those that did not.

**Table A1:** Comparison of demographics between completers and non-completers.

| Participants, *n* (% of total enrolled) | Coaches who completed intervention | Coaches lost to follow up or non-completers |
| --- | --- | --- |
| Total, *n* (% of number enrolled) | 27 (53%) | 24 (47%) |
| Pronouns, *n* (%) |  |  |
| She/her | 22 (43%) | 19 (37%) |
| He/him | 4 (8%) | 5 (10%) |
| They/them | 1 (2%) | 0 (0%) |
| Age (years), mean (SD) | 34 (9.5) | 35 (9.3) |
| Range | 17-57 | 18-47 |
| Location, *n* (%) |  |  |
| Victoria | 10 (20%) | 9 (18%) |
| Queensland | 4 (8%) | 4 (8%) |
| New South Wales | 4 (8%) | 3 (6%) |
| Australian Capital Territory | 3 (6%) | 2 (4%) |
| South Australia | 3 (6%) | 2 (4%) |
| Western Australia | 2 (4%) | 2 (4%) |
| Tasmania | 1 (2%) | 2 (4%) |
| Years of experience teaching circus to 3-5year old children, *n* (%) |  |  |
| <1 | 3 (6%) | 1 (2%) |
| 1-5 | 11 (22%) | 9 (18%) |
| 6-10 | 6 (12%) | 7 (14%) |
| 11-15 | 2 (4%) | 2 (4%) |
| 16-20 | 3 (6%) | 3 (6%) |
| >20 | 2 (4%) | 2 (4%) |

## Appendix B: Theoretical Framework of Acceptability (TFA) questionnaire results

**Figure B1:**
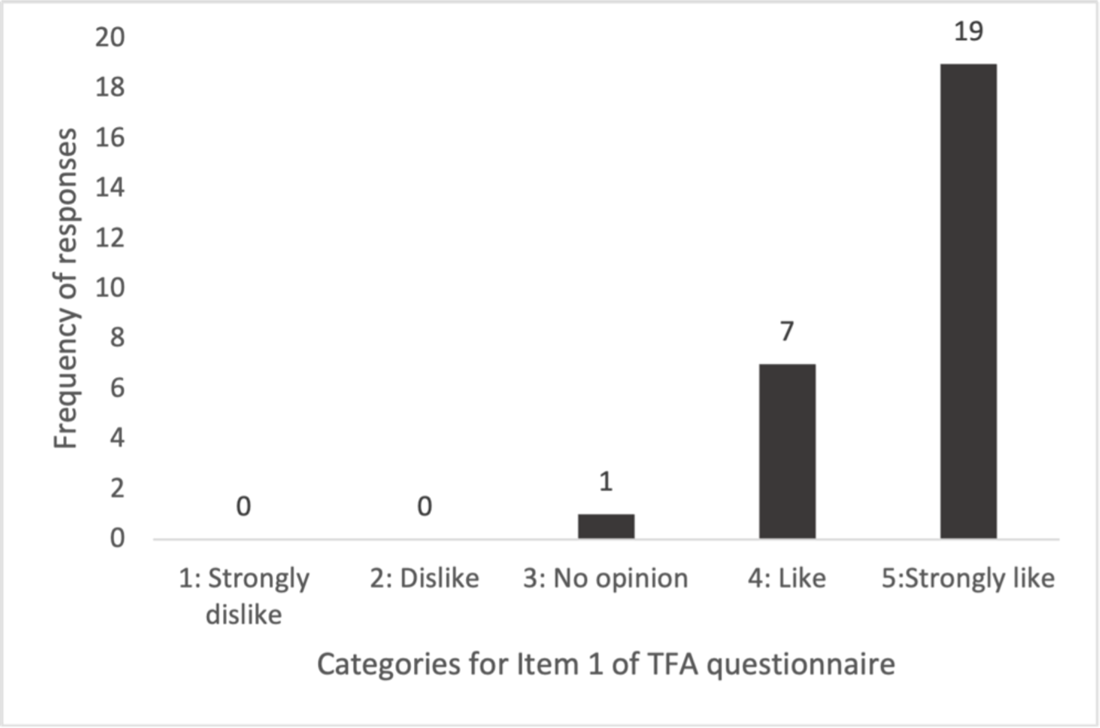
Participant responses to the question *“Did you like or dislike the Coach Training program?”*: Item 1 of the TFA questionnaire, Affective Attitude [34].

**Figure B2:**
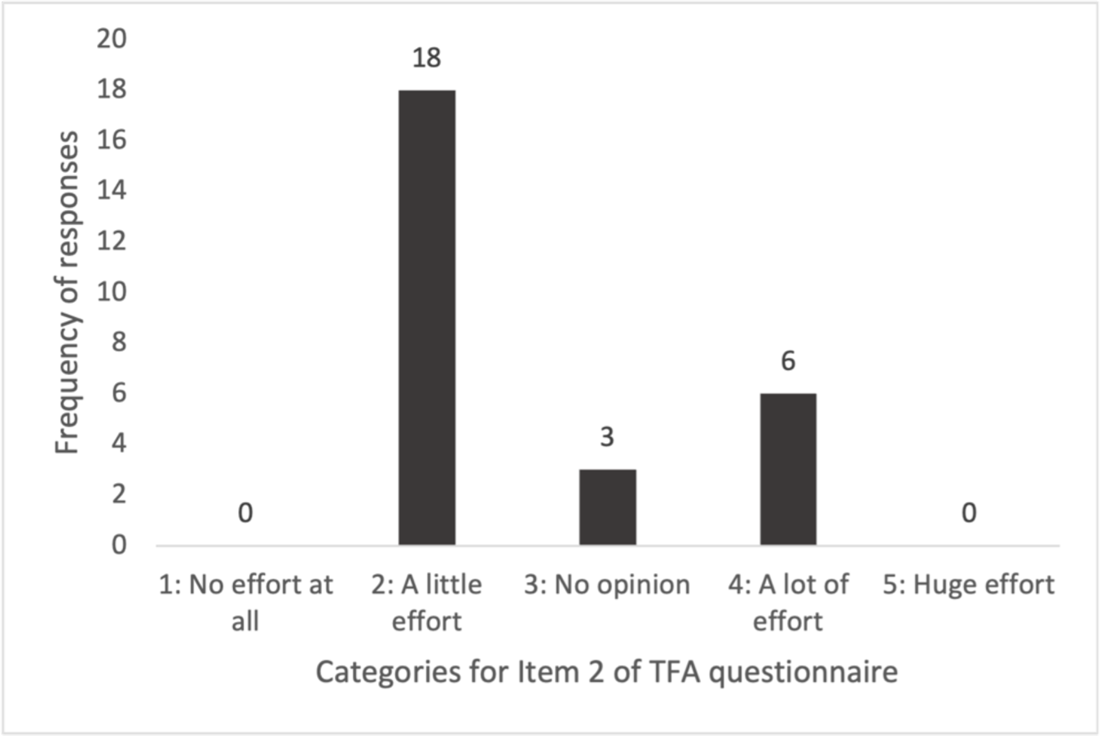
Participant responses to the question *“How much effort did it take to participate in the Coach Training?”*: Item 2 of the TFA questionnaire, Burden [34].

**Figure B3:**
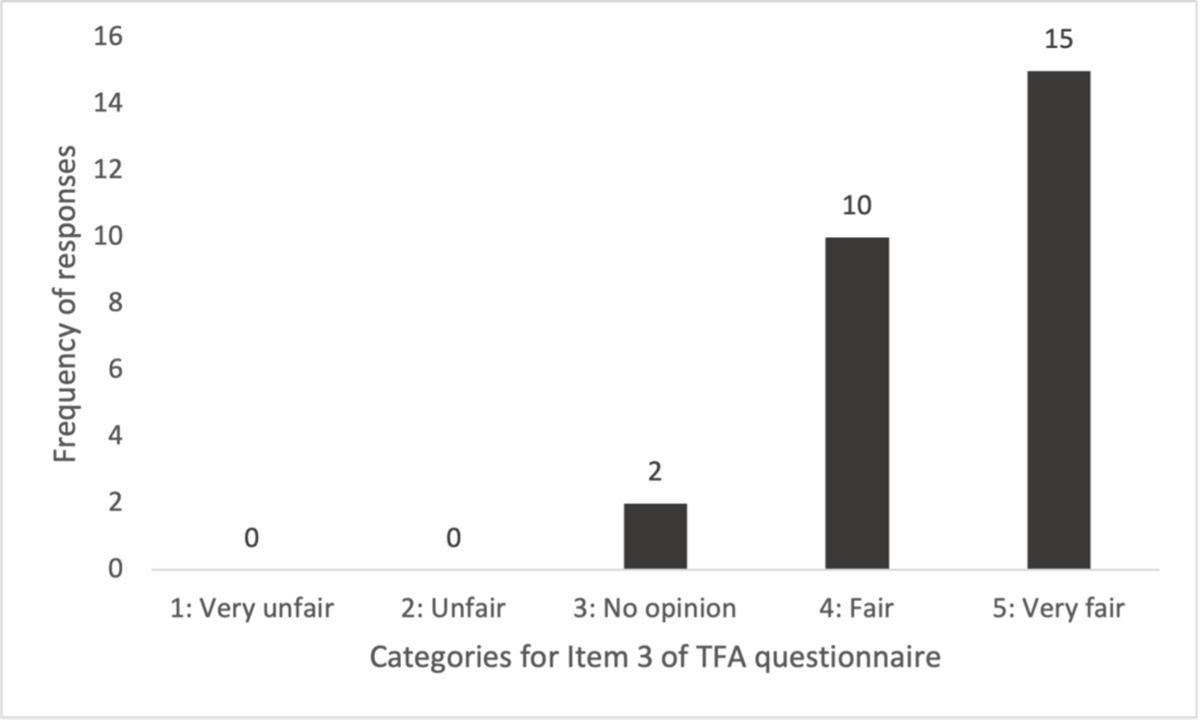
Participant responses to the question *“How fair is the Coach Training program for circus coaches?”:* Item 3 of the TFA questionnaire, Ethicality [34].

**Figure B4:**
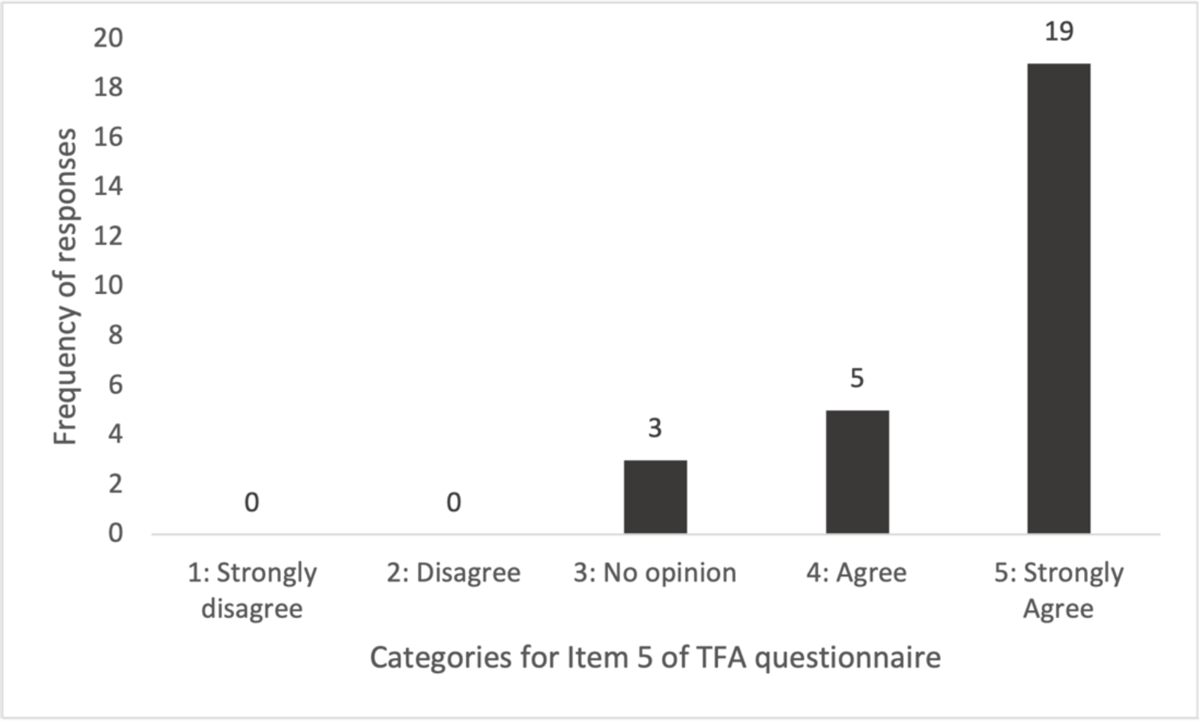
Participant responses to the question *“The Coach Training has improved my knowledge & confidence in working with children born preterm?”:* Item 4 of the TFA questionnaire, Perceived Effectiveness [34].

**Figure B5:**
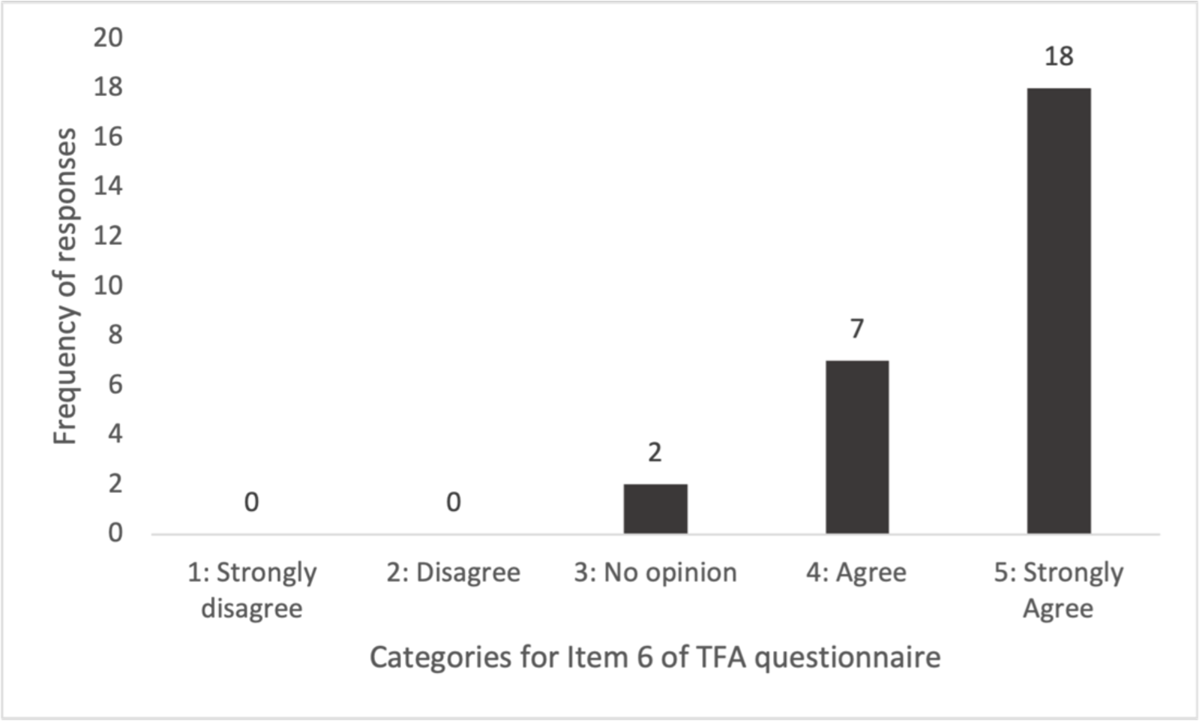
Participant responses to the statement *“It is clear to me how the Coach Training improves coaches’ knowledge & confidence in working with children born preterm”:* Item 5 of the TFA questionnaire [34].

**Figure B6:**
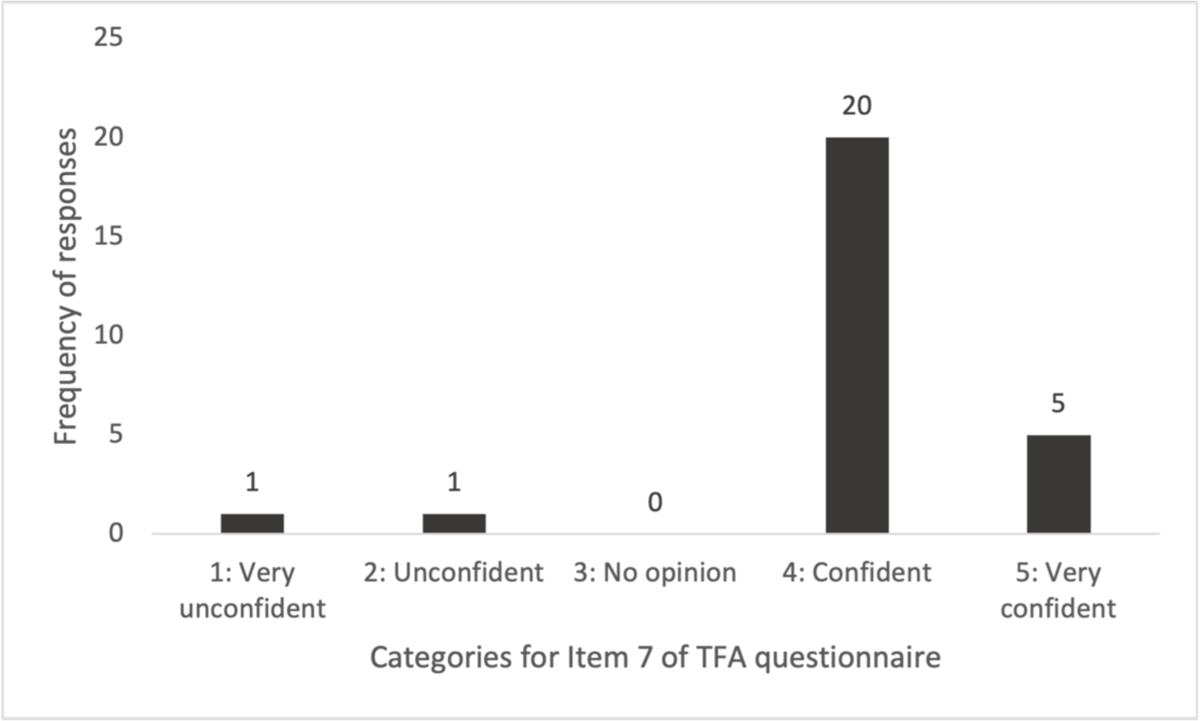
Participant responses to the question *“How confident did you feel about participating in the Coach Training?”*: Item 6 of the TFA questionnaire [34].

**Figure B7:**
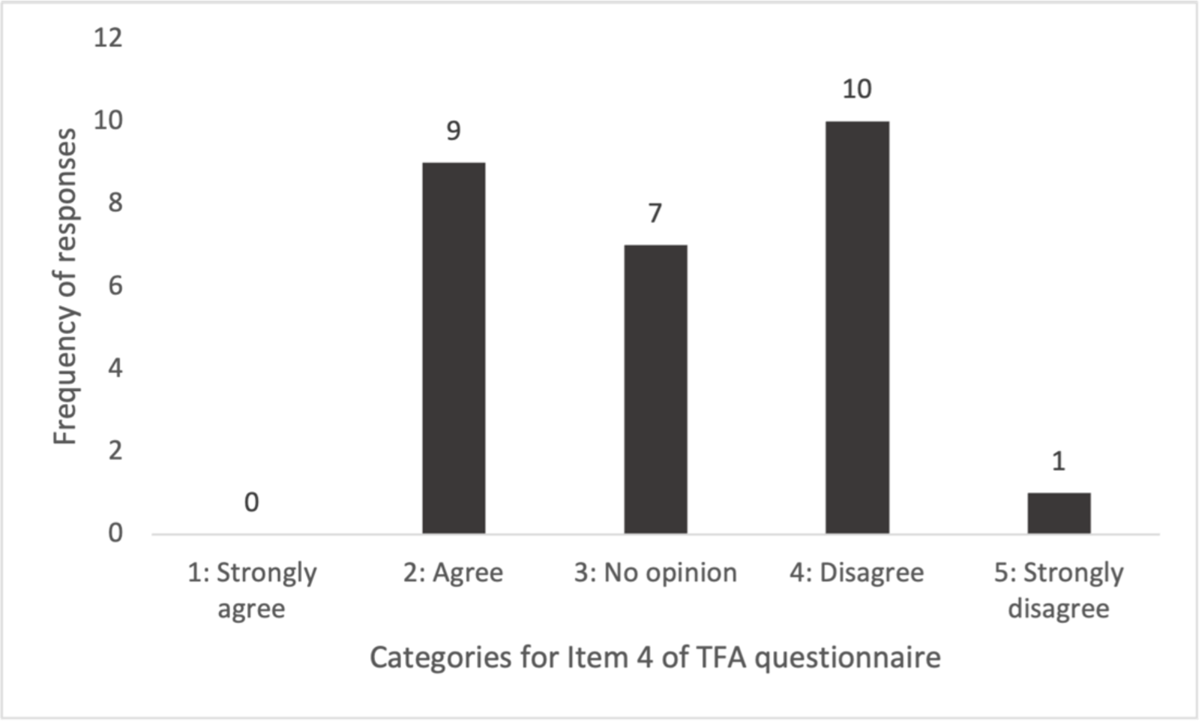
Participant responses to the statement *“Completing the Coach Training interfered with my other priorities”:* Item 7 of the TFA questionnaire, Opportunity Costs [34].

## Appendix 3: Determinants of Implementation Behaviour Questionnaire (DIBQ) results

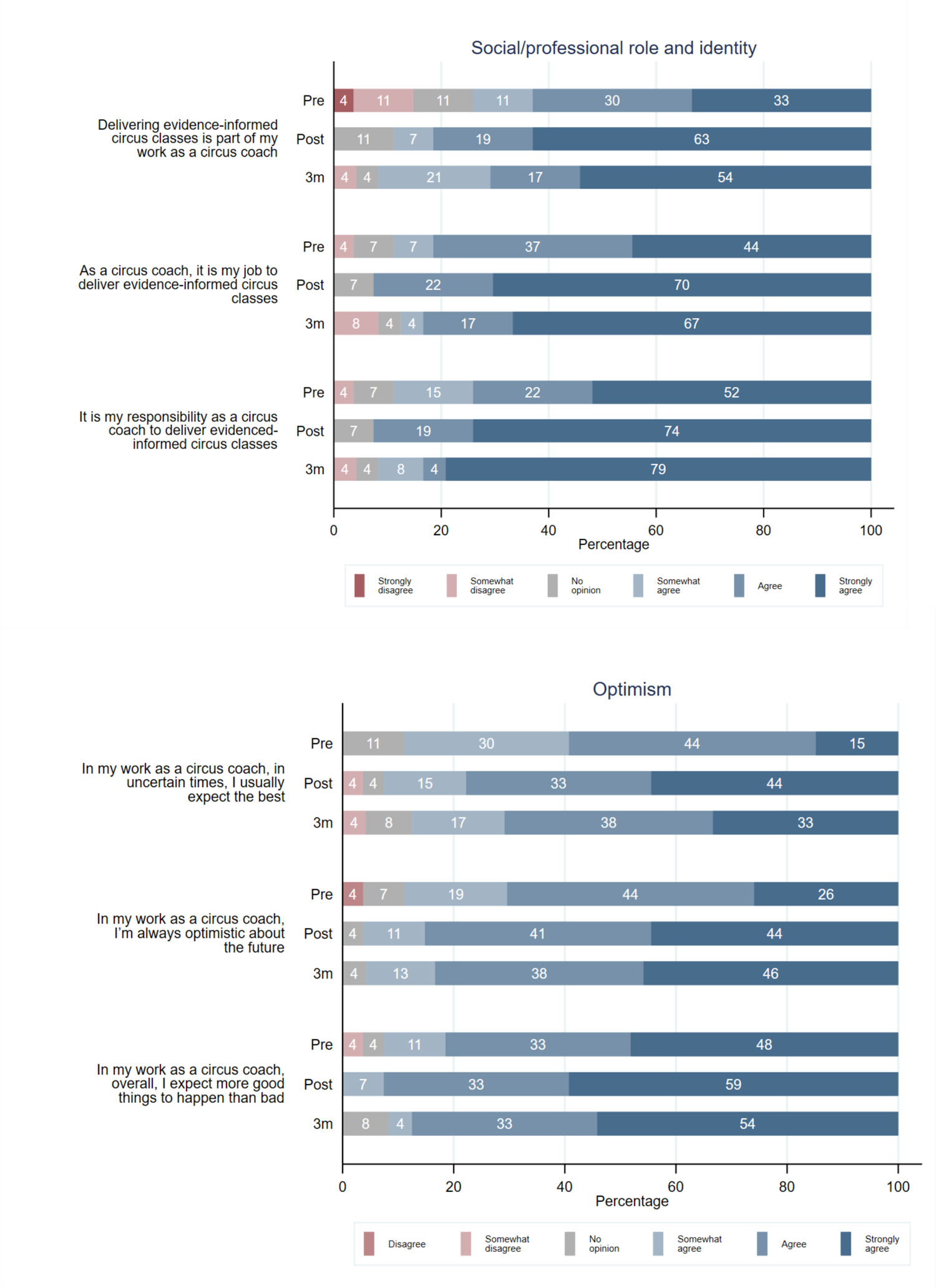
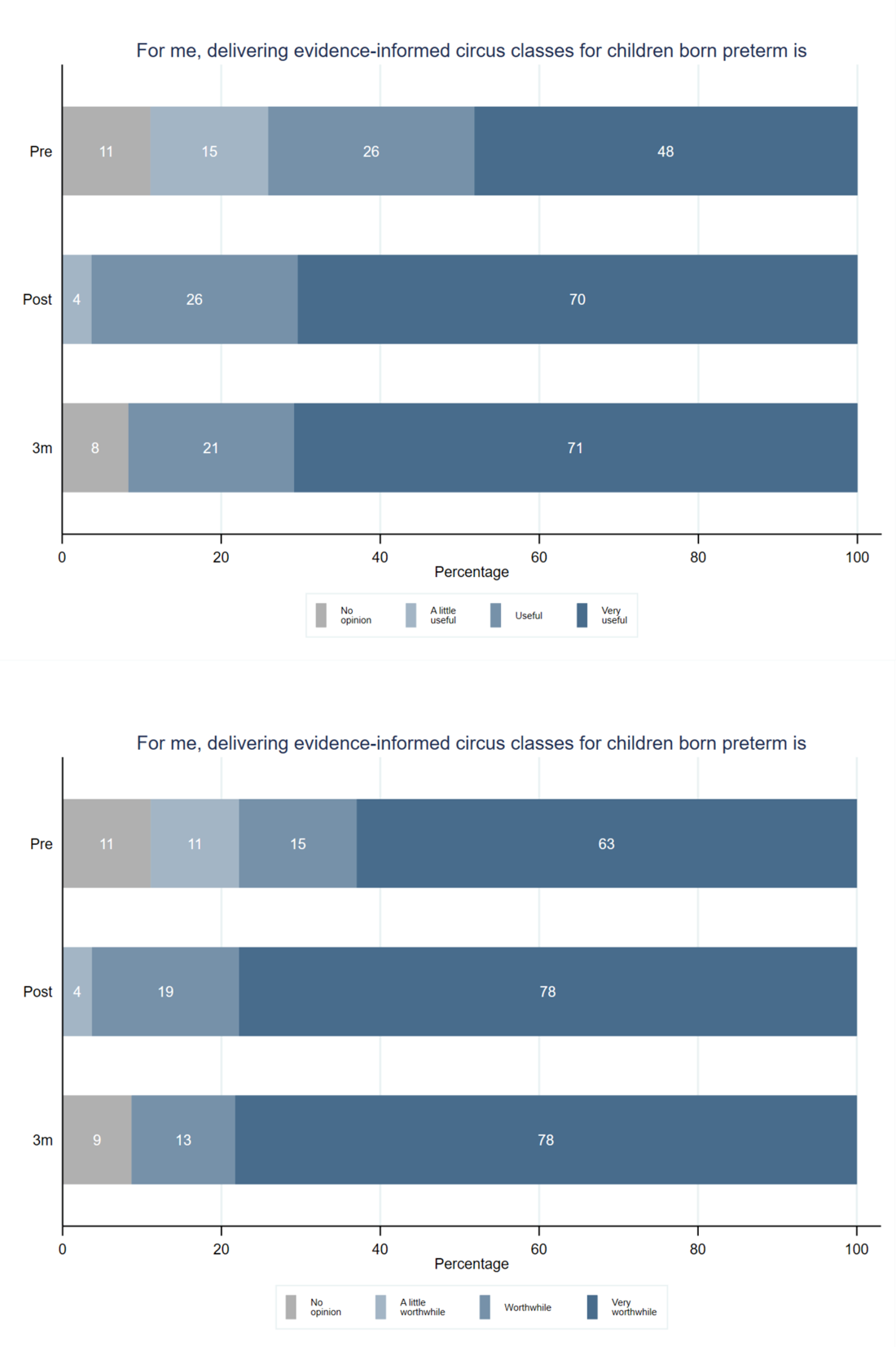
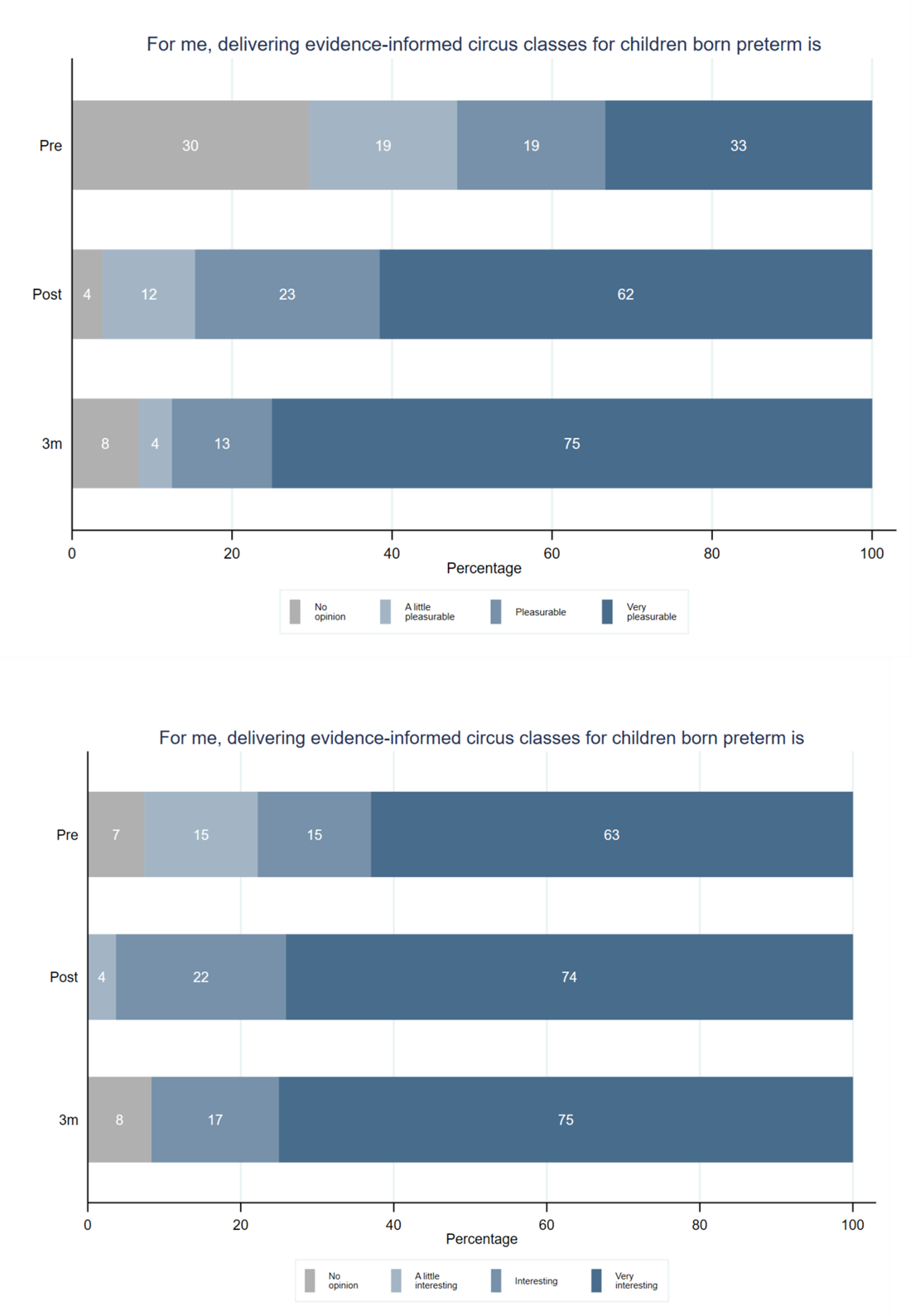
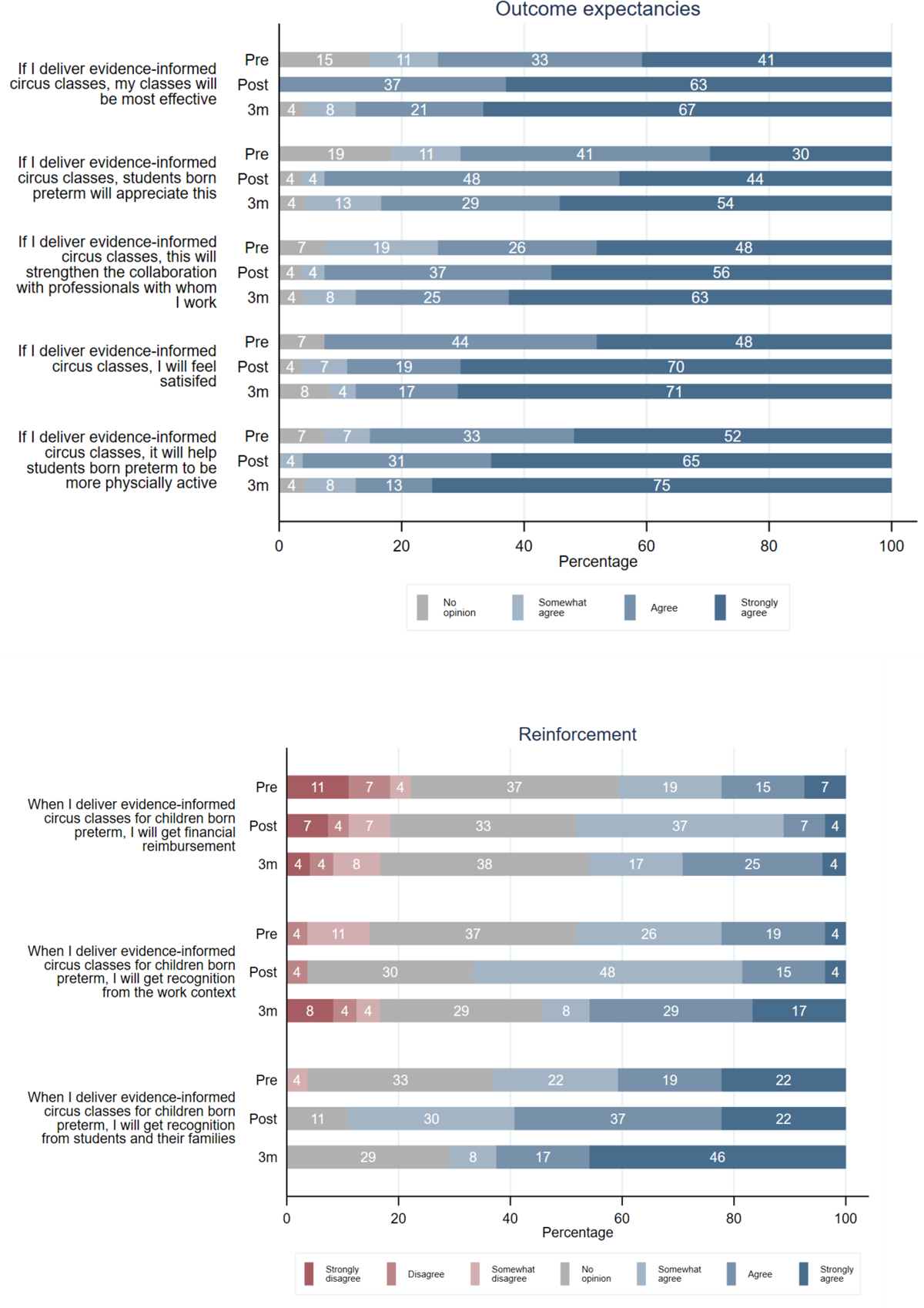
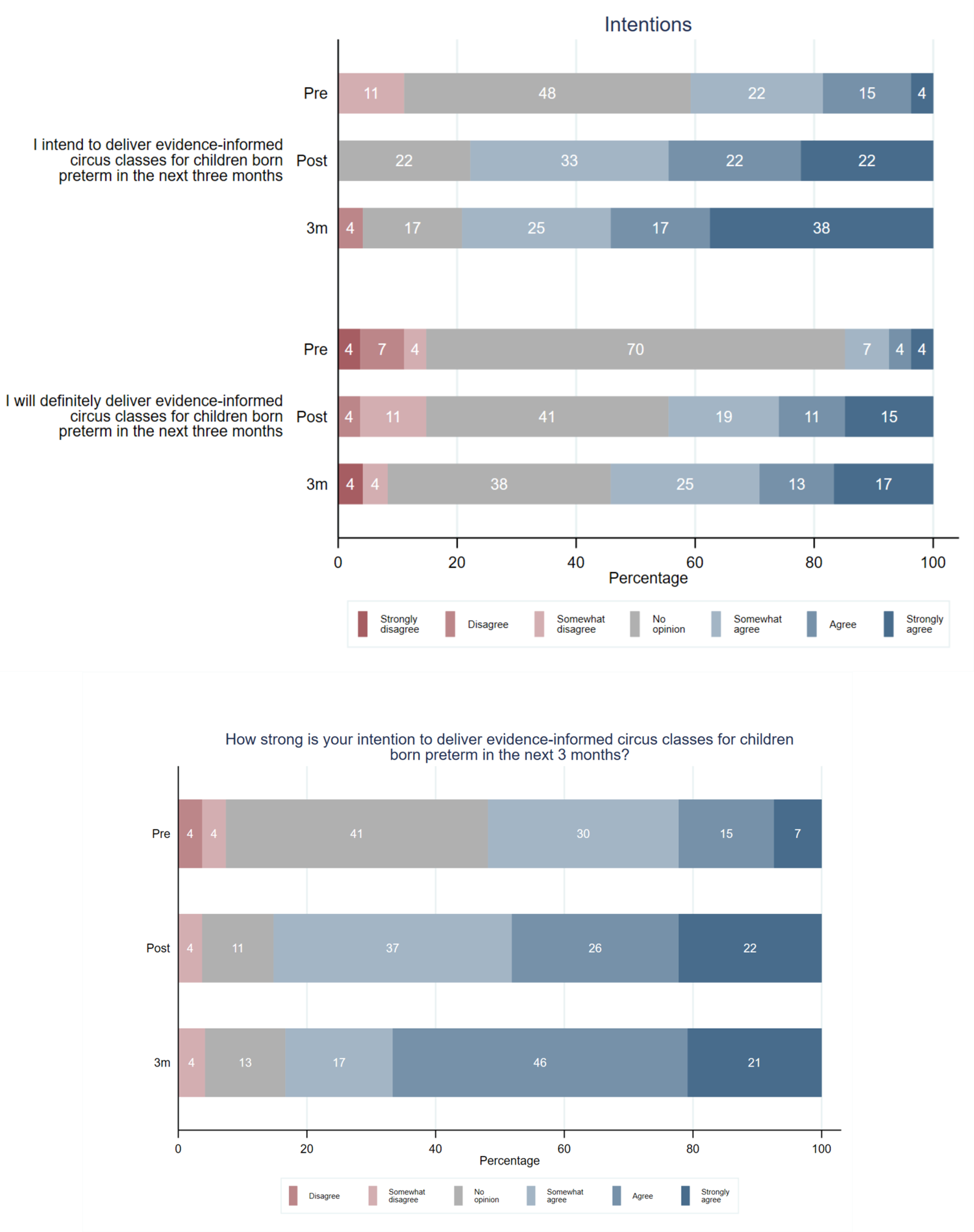
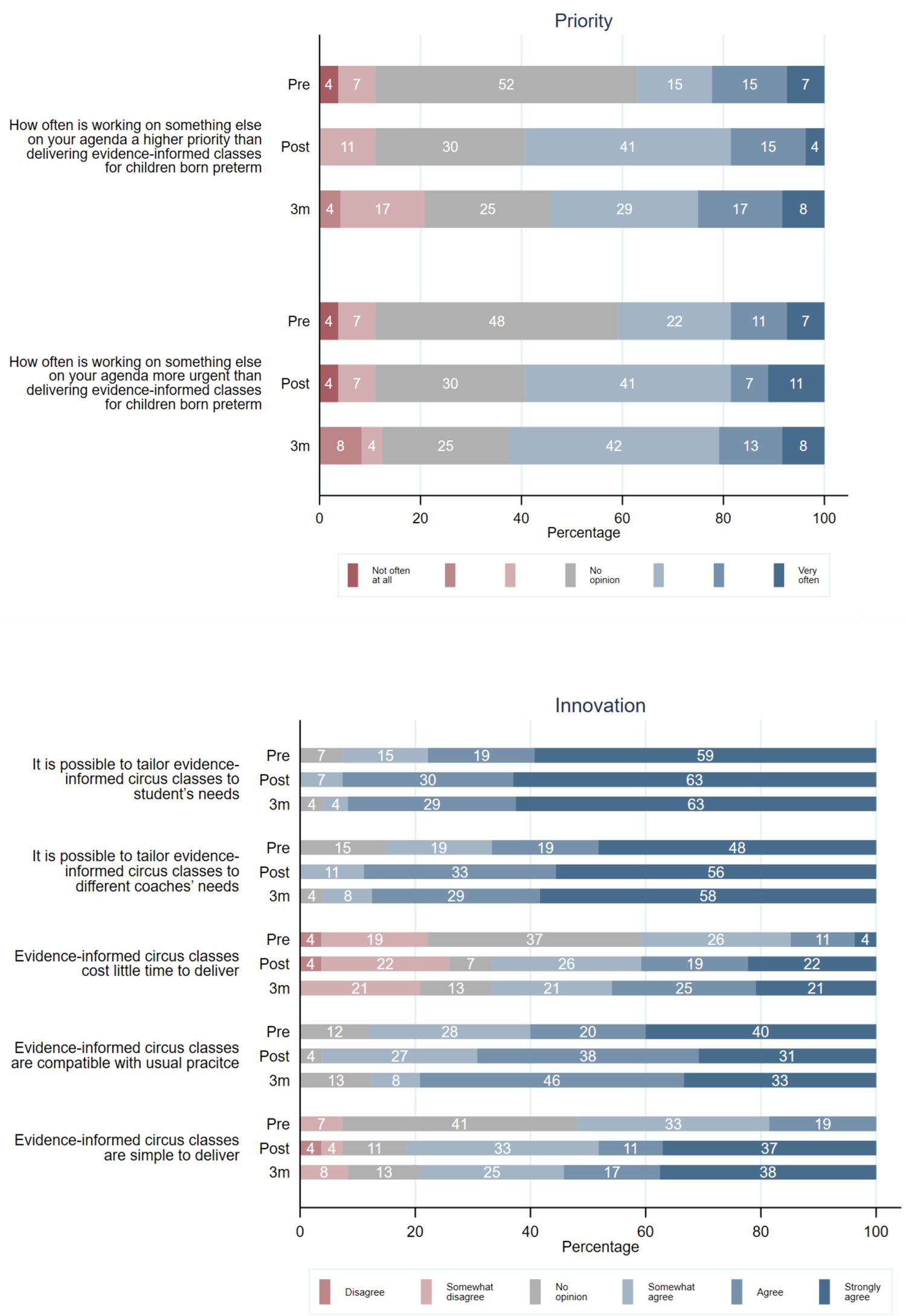
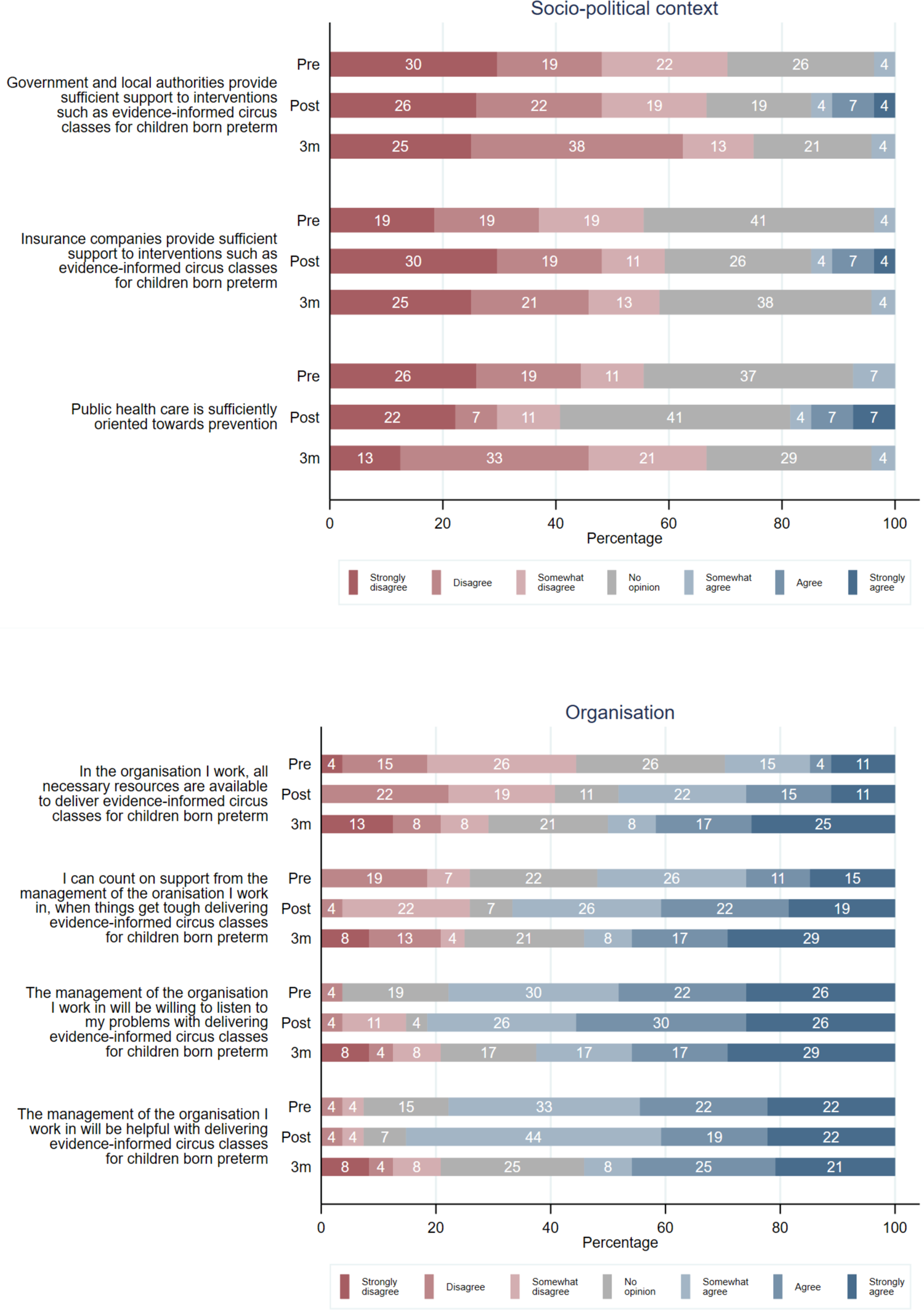
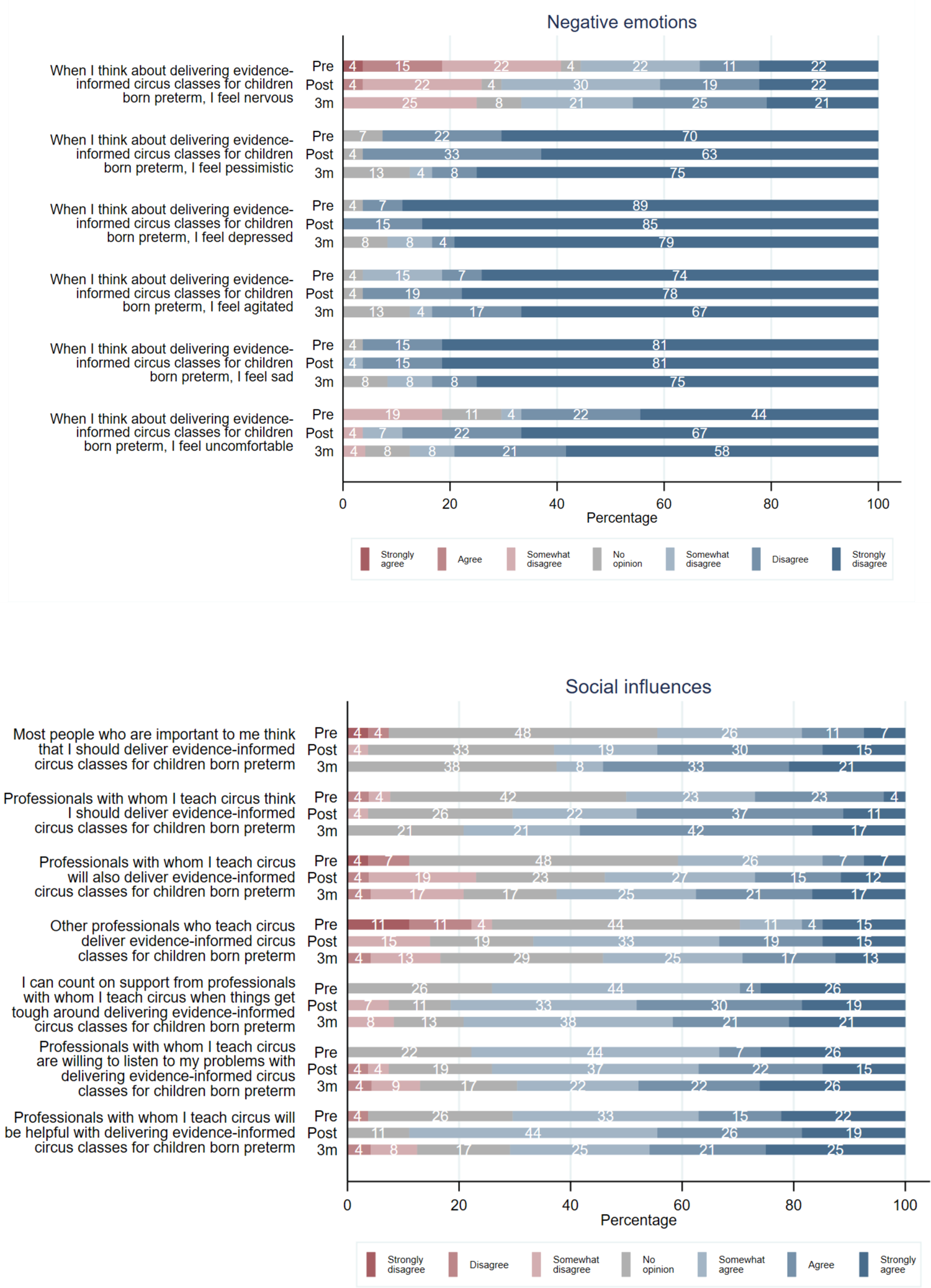
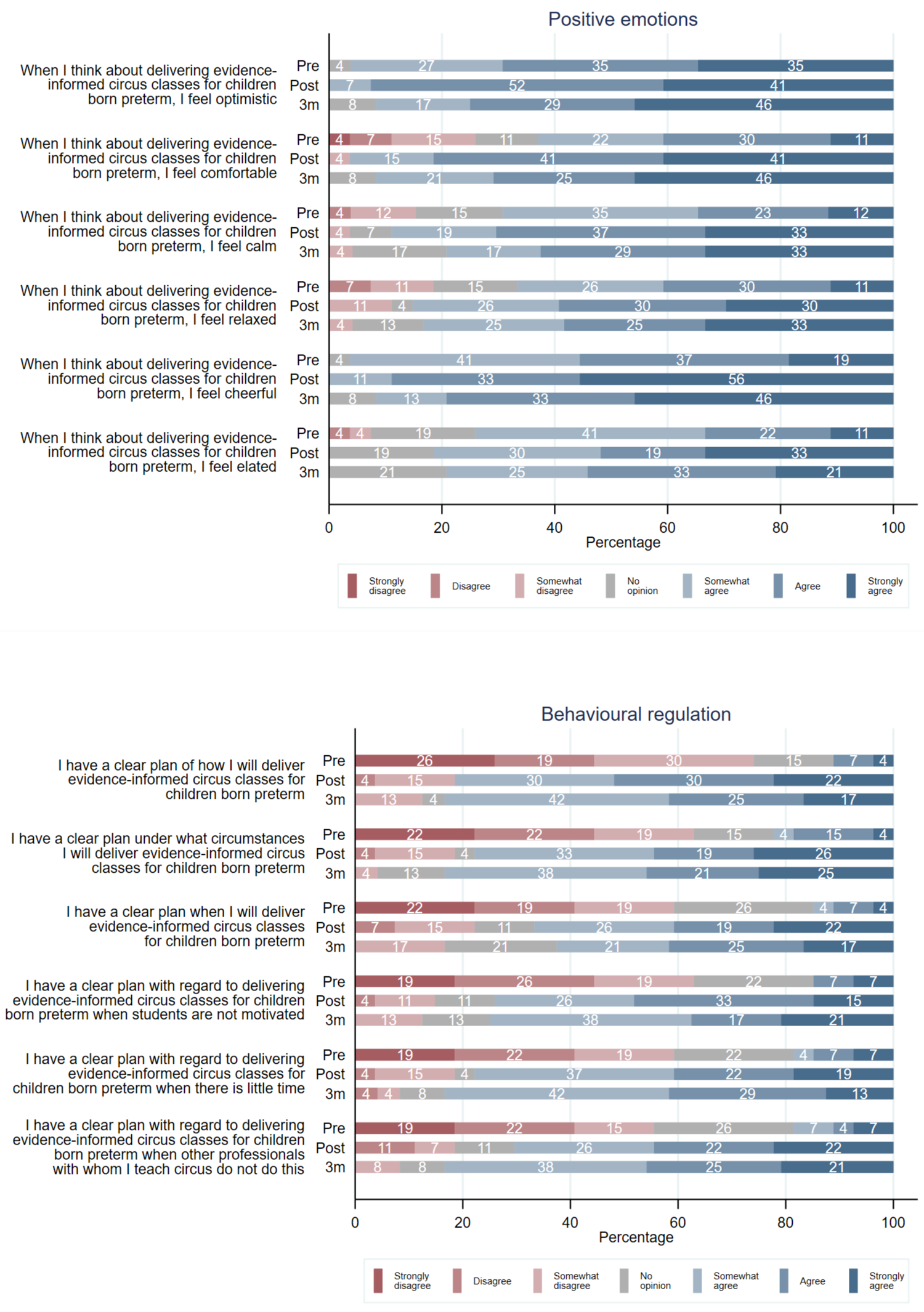
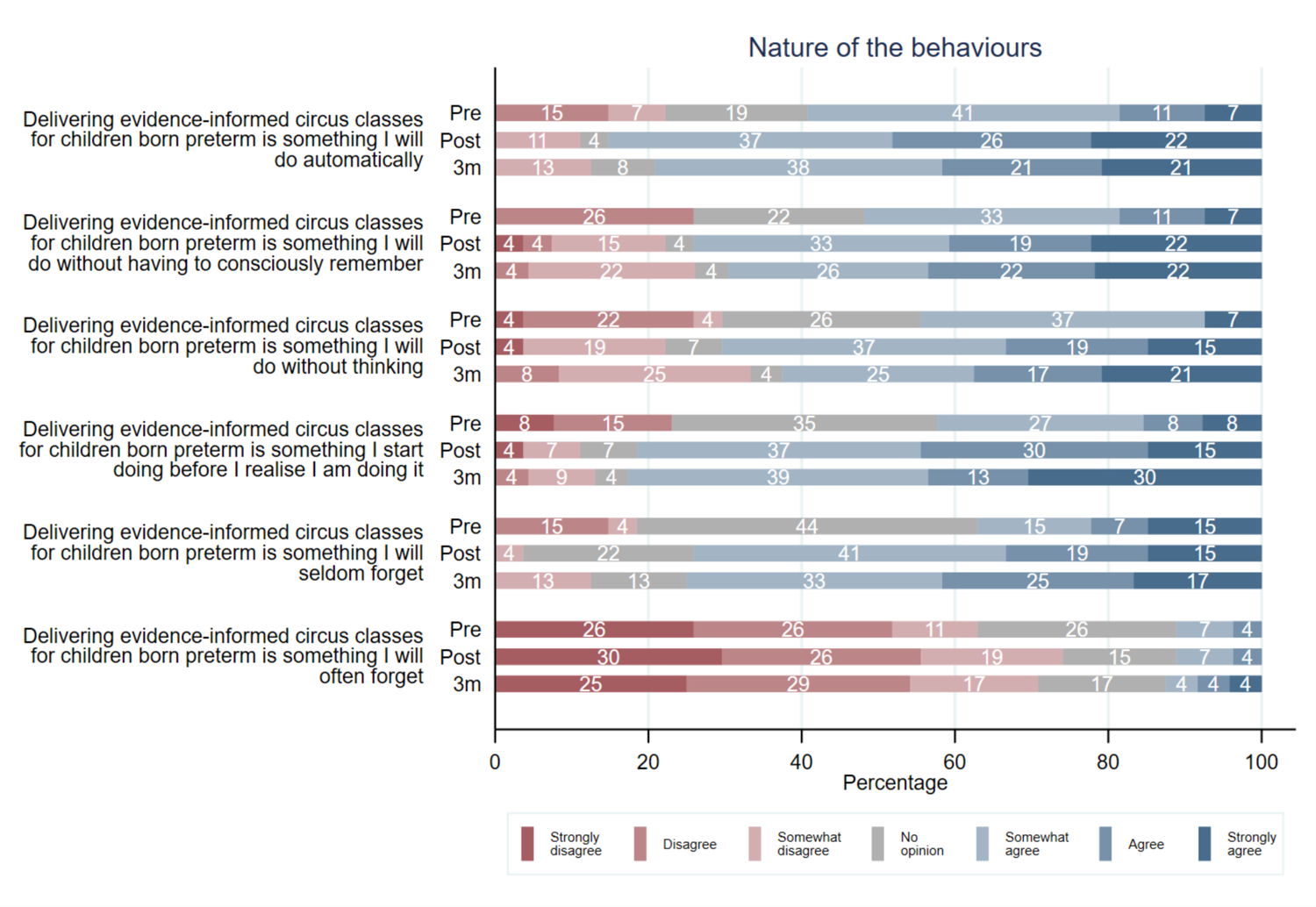

